# Geospatial patterns and predictors of neonatal mortality among HIV infected and non-infected mothers of rural Zambia: A comparative analysis of the 2018 Zambia Demographic and Health Survey

**DOI:** 10.1101/2024.05.22.24307735

**Authors:** Samson Shumba, Deborah Tembo, Miyanda Simwaka, Nedah Chikonde Musonda, Chipo Nkwemu, Sebean Mayimbo

## Abstract

Neonatal mortality is the death of a live-born infant within the first 28 completed days of life. Neonatal mortality remains a major public health concern in most African countries, with the Sub-Saharan region being the highest contributor at 27 deaths per 1000 live births, accounting for 43% of the total number of deaths. Zambia still fares poorly in terms of neonatal mortality, ranking 162 out of 195 countries globally. The study aimed to investigate the spatial patterns and predictors of neonatal mortality in rural Zambia. This study utilized the national-level data from the Zambia Demographic and Health Survey (ZDHS) program, utilizing the survey dataset from 2018. Statistical analyses were conducted using the Rao – Scott Chi-square test to assess associations between dependent and independent variables. Additionally, a multilevel mixed effect logistic regression model was used to examine predictors of neonatal mortality. Geospatial patterns of neonatal mortality across Zambia’s ten provinces were investigated using Quantum Geographical Information System (QGIS) version 3.34.1 to generate a univariate choropleth map. Data analysis was performed using Stata version 14.2. The study findings indicate a higher mortality rate among HIV-infected mothers aged 45 to 49 (100%) compared to 95.21% among non-HIV-infected mothers. Neonatal mortality was notably elevated among neonates born to mothers with no education (40.78%) and those with HIV infection (54.56%). Employment status also influenced mortality rates, with employed non-HIV-infected mothers showing 36.35% mortality compared to 49.39% among unemployed HIV-infected mothers. Higher birth weights, particularly 4000 grams or higher, were associated with increased mortality for both HIV-infected (81.15%) and non-infected (21.00%) mothers. Multilevel logistic regression identified predictors of mortality, including maternal age (40 to 44 years), neonate’s sex (female), and maternal HIV status. Geospatial analysis revealed Eastern and North-Western provinces as hotspots for neonatal mortality among HIV-infected mothers, while Muchinga was a hotspot for non-HIV-infected mothers. This study examined neonatal mortality among HIV-infected and non-infected mothers in rural Zambia, uncovering predictors such as maternal age, neonate sex, birthweight, maternal HIV status, and community desired number of children. Regional variations in mortality underscore the necessity for targeted interventions. Strengthening healthcare infrastructure, community outreach, healthcare worker training, maternal education, and addressing regional differences are crucial for improving maternal and child health and meeting Sustainable Development Goals targets.

## Introduction

Neonatal mortality is characterized by the death of a live-born infant, irrespective of the gestational age at birth, within the initial 28 completed days of life [1]. The initial month after birth represents a critical phase for child survival, marked by the unfortunate loss of 2.4 million newborns globally in the year 2020 [2]. During that same year, nearly 47% of all deaths in children under the age of five occurred during the neonatal period, which spans the first 28 days of life and 98% of the deaths occurred in low and middle-income countries (LMIC) [1–3]. This percentage reflects a notable increase from the 40% reported in 1990. The world has made impressive progress towards the survival of children since 1990. In 2020, the number of neonatal deaths declined from 5 million in 1990 to 2.4 million, nonetheless, the decline in neonatal mortality from 1990 to 2020 has been slower than that of the post-neonatal under-five mortality [1].

Sub-Saharan Africa bears the highest global neonatal mortality rate, reaching approximately 27 deaths per 1000 live births, and plays a significant role in global newborn fatalities, accounting for 43% of the total [4,5]. In close proximity, central and southern Asia report a neonatal mortality rate of 23 deaths per 1000 live births, contributing to 36% of global newborn deaths [1]. Moreover, a child born in Sub-Saharan Africa faces a staggering tenfold higher likelihood of dying in the first month than a child born in a high-income country. In Uganda a study revealed that the underlying causes of death are related to poor access and low utilization of health services during pregnancy and childbirth [6]. Furthermore, findings have revealed that more newborn deaths occur at home among the rural poor [7].

Zambia relative to other countries fairs poorly ranking 162 out of 195 countries globally in terms of neonatal mortality in Zambia [8]. According to Zambia Statistical Agency (2018) it indicated that neonatal mortality rate decreased from 37 to 27 neonatal deaths per 1000 live births between 2001 and 2018 [9]. Moreover, various studies have identified several factors correlated with heightened neonatal mortality rates in both the general population and facility-based settings. These include lower levels of maternal education [10–12], place of delivery and residence [13,14], maternal age [15], parity [16], gestational age [10,15], inadequate antenatal visits [17], newborn sex [13], newborn age [18], low birth weight [19], maternal and fetal complications [18], preterm birth [19], and hypothermia [20]. Despite these findings, there remains a gap in understanding neonatal mortality and its associated risk factors, particularly within rural communities, especially between HIV-infected and non-infected mothers in rural Zambia. Therefore, this study aimed to investigate the geospatial patterns and predictors of neonatal mortality among both HIV-infected and non-infected mothers in rural Zambia.

## Methods

This study constitutes a secondary analysis of microdata utilizing national-level data sourced from the Zambia Demographic and Health Survey (ZDHS) program using the survey dataset in 2018. The ZDHS is a comprehensive, nationally representative household survey conducted by the Zambia Statistics Agency in collaboration with global partners, including ICF International and the United States Agency for International Development (USAID). The survey employs a two-stage sampling process, initially selecting enumeration areas (EAs) and subsequently households.

### Dependent and independent variables

The variable of interest in this study is neonatal mortality (that is death of babies between 0 and 28 days). The explanatory variables that were used in the study were demographic, socio-economic, behavioral, and community-level factors. These variables included the following: the mother’s age (categorized as 15-19, 20-24, 25-29, 30-34, 35-39, 40-44, or 45-49 years), current marital status of the mother (categorized as not married or married), the residence of the mother (categorized as urban or rural), education level of the mother (classified as no education, primary, secondary, tertiary), employment status of the mother (categorized as employed or unemployed), and household wealth index of the mother (categorized as poorest, poorer, middle, richer, or richest). The household wealth index is computed based on readily available data concerning a household’s possession of selected assets, such as televisions and bicycles; materials utilized for housing construction; and types of water access and sanitation facilities [21].

### Community level variables

Community-level variables in this study were derived by aggregating individual-level data into clusters and incorporated community poverty, community education, community knowledge of family planning (FP) methods, and place of residence. These community-level variables were dichotomized as either ’low’ or ’high,’ reflecting the extent of the phenomena under investigation at the cluster level. Place of residence and geographical region were maintained in their original categorizations. Place of residence played a pivotal role in the sample design, as it was utilized as a criterion to estimate the prevalence of key demographic and health indicators at the national level. It was categorized as either ’rural’ or ’urban’ and directly contributed to the description of community characteristics.

### Data Analysis

For descriptive purposes, frequencies and percentages were computed for categorical variables. To determine the association between the outcome variable (neonatal mortality) and the categorical variables, the Uncorrelated Design Based Chi-square test (Rao – Scott Chi-square test) was used. The multilevel mixed-effect logistic regression model was employed to determine the factors affecting newborn mortality. Additionally, the study used an investigator-led approach, all variables were selected from a wide range of literature. The Intra-Class Correlation (ICC), Akaike Information Criteria (AIC), and the Bayesian Information Criteria (BIC) were sufficiently explored to select the best-fit model. Stata version 14.2 was used for analysis.

### Geospatial Analysis

To examine the clustering and spatial distribution of neonatal mortality across provinces in Zambia, this study utilized Quantum Geographic Information System (QGIS) version 3.34.1 to construct a univariate choropleth map. Spatial analysis was conducted at the provincial level, with each neonate matched to their provincial residence using geo-coordinate data collected from the Demographic and Health Survey (DHS). The DHS employs pre-defined data assigning each case to a province. Spatial analysis units were defined as clusters of sample households as designated by the Zambian Demographic and Health Survey (ZDHS). The coordinate system utilized was the World Geographic System (WGS) 1984 Universal Transverse Mercator (UTM) Zone 36S.

### Ethics statement

The methodologies used in the 2018 ZDHS, including biomarker measurement protocols, were ethically cleared by both the Inner City Fund (ICF) institutional review boards (IRBs) and the Tropical Diseases Research Centre (TDRC) in Zambia. Oral consent was obtained from respondents, with parental or guardian consent for adolescents under 18. Neonate information was collected from caregivers. Detailed information on the DHS consent process is available (https://www.dhsprogram.com/What-We-Do/Protecting-the-Privacy-of-DHS-Survey-Respondents.cfm). Authorization to use ZDHS data was granted by ICF Macro, and the dataset can be accessed (https://www.dhsprogram.com/data). The user strictly adhered to confidentiality guidelines, ensuring the anonymity of respondents.

## Results

Participation in the survey was limited to children aged one month from selected households of women who had consented to take part in the research. Detailed methods employed in the DHS are comprehensively documented elsewhere [9]. A total of 2015 participants were enrolled into the study comprising neonates from HIV-infected and non-infected mothers as shown in Fig 1.

**Figure 1:**
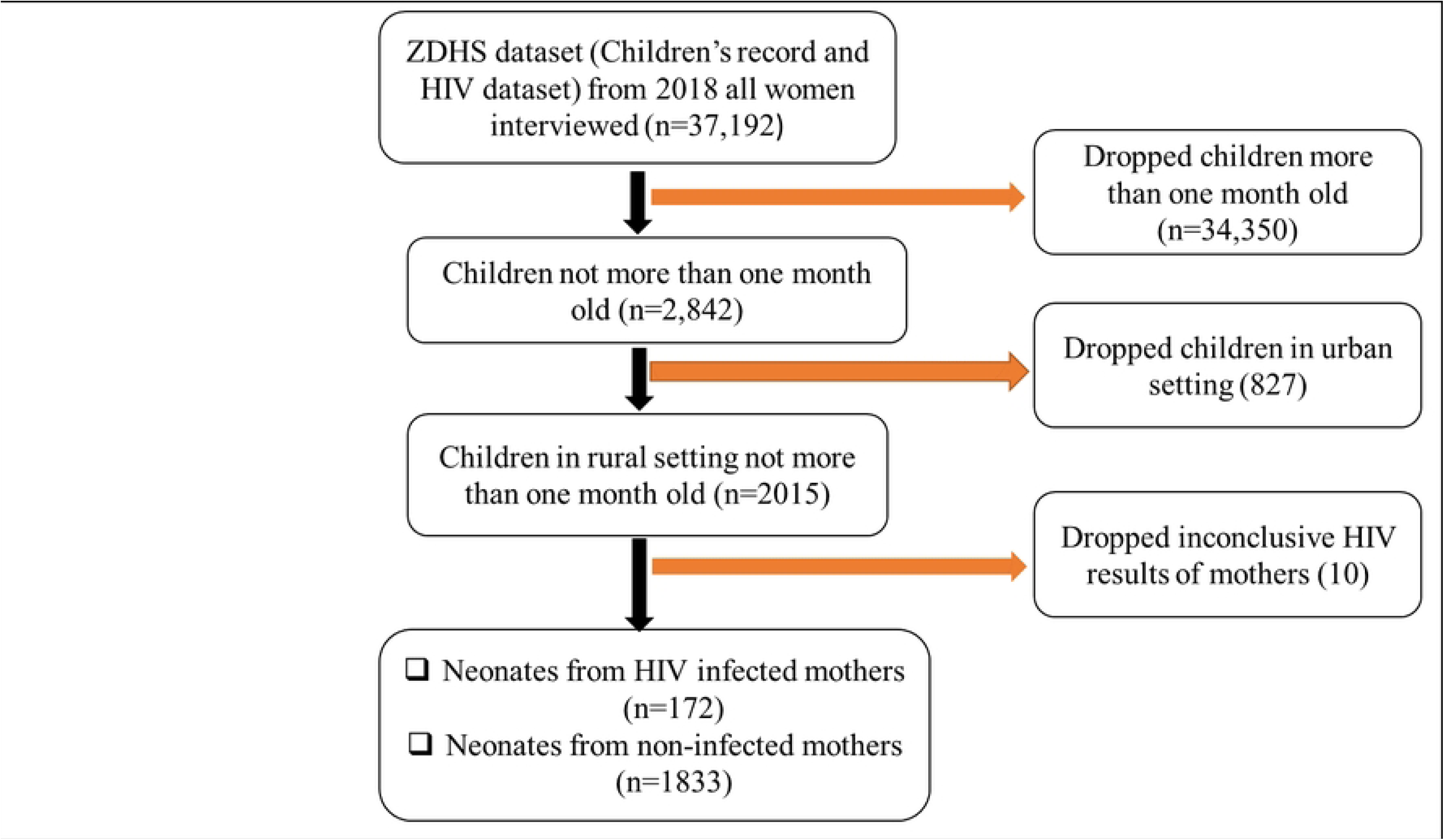
Description of sample derivation criteria. (Source: Author generated)

The data presented in Table 2 provides an overview of neonatal mortality in rural settings among mothers infected with HIV and those not infected with HIV, drawing from the 2018 Zambia Demographic and Health Survey dataset. The findings reveal distinct patterns in neonatal mortality across various background characteristics, including maternal age, education level, employment status, neonate gender, birth order, birthweight, place of delivery, and HIV status. Notably, neonatal mortality increases significantly with maternal age (p<0.0001), with the highest mortality observed in the age group 45 to 49 (100%) among HIV-infected mothers compared to 95.21% in non-HIV infected mothers.

**Table 2:**
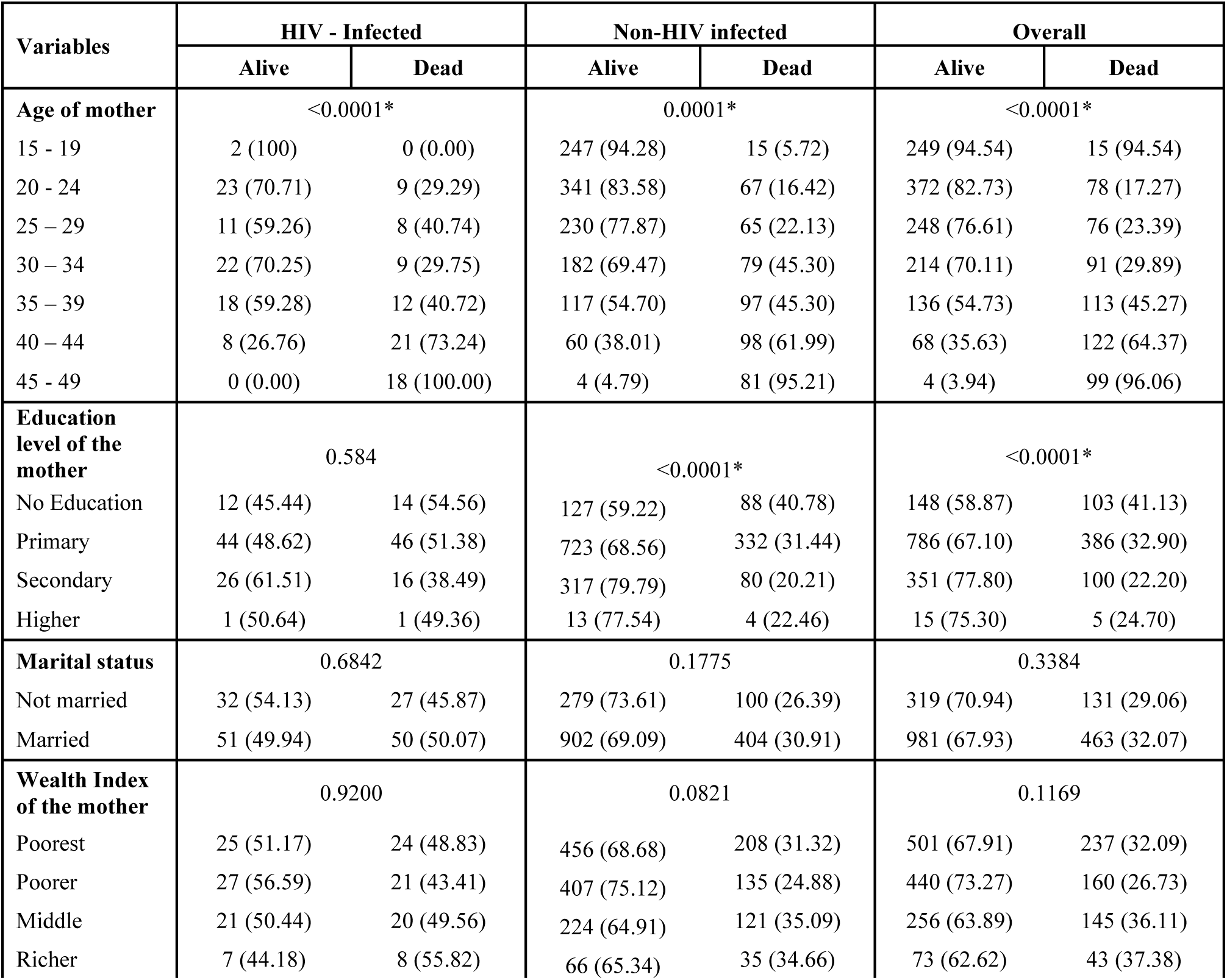
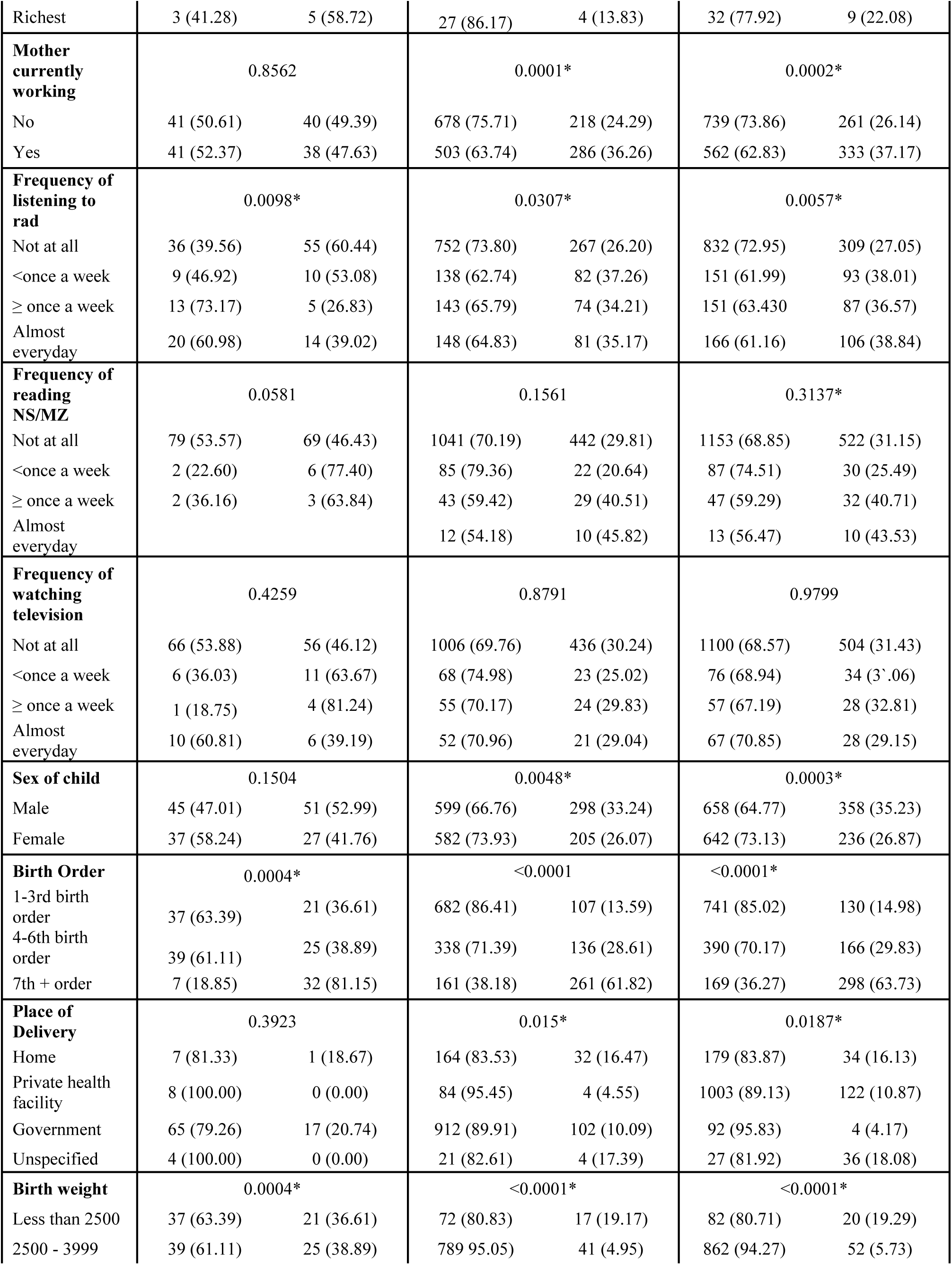
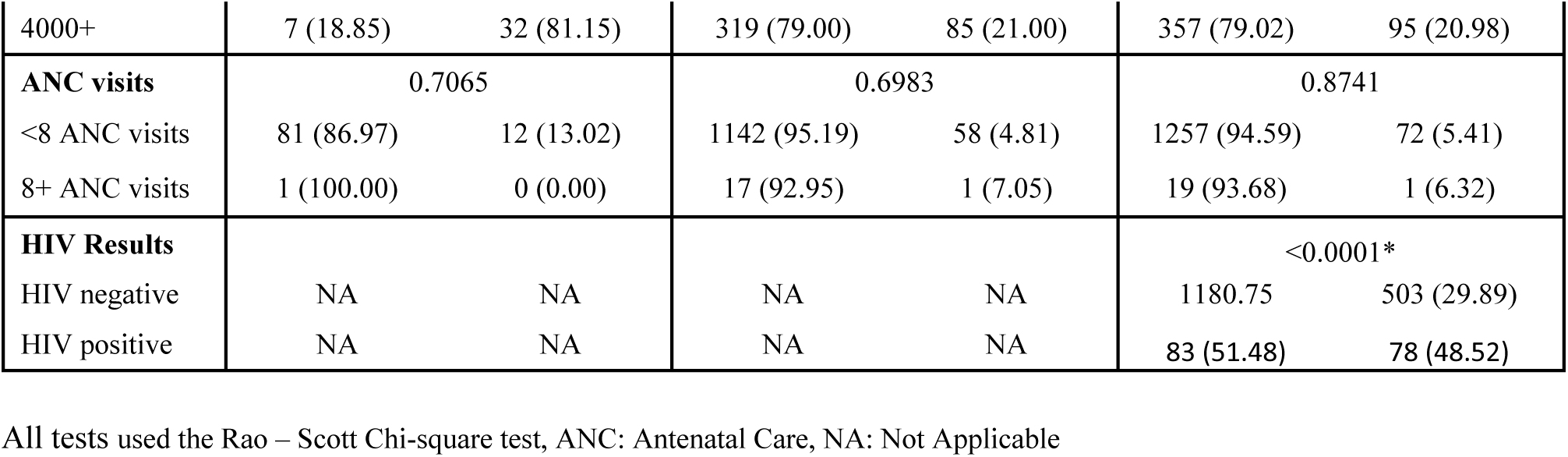
Key attributes of neonatal mortality in rural setting of Zambia in 2018.

| Variables | HIV - Infected |  | Non-HIV infected |  | Overall |  |
| --- | --- | --- | --- | --- | --- | --- |
|  | Alive | Dead | Alive | Dead | Alive | Dead |
| **Age of mother** | <0.0001* |  | 0.0001* |  | <0.0001* |  |
| 15 - 19 | 2 (100) | 0 (0.00) | 247 (94.28) | 15 (5.72) | 249 (94.54) | 15 (94.54) |
| 20 - 24 | 23 (70.71) | 9 (29.29) | 341 (83.58) | 67 (16.42) | 372 (82.73) | 78 (17.27) |
| 25 - 29 | 11 (59.26) | 8 (40.74) | 230 (77.87) | 65 (22.13) | 248 (76.61) | 76 (23.39) |
| 30 - 34 | 22 (70.25) | 9 (29.75) | 182 (69.47) | 79 (45.30) | 214 (70.11) | 91 (29.89) |
| 35 - 39 | 18 (59.28) | 12 (40.72) | 117 (54.70) | 97 (45.30) | 136 (54.73) | 113 (45.27) |
| 40 - 44 | 8 (26.76) | 21 (73.24) | 60 (38.01) | 98 (61.99) | 68 (35.63) | 122 (64.37) |
| 45 - 49 | 0 (0.00) | 18 (100.00) | 4 (4.79) | 81 (95.21) | 4 (3.94) | 99 (96.06) |
| **Education level of the mother** | 0.584 |  | <0.0001* |  | <0.0001* |  |
| No Education | 12 (45.44) | 14 (54.56) | 127 (59.22) | 88 (40.78) | 148 (58.87) | 103 (41.13) |
| Primary | 44 (48.62) | 46 (51.38) | 723 (68.56) | 332 (31.44) | 786 (67.10) | 386 (32.90) |
| Secondary | 26 (61.51) | 16 (38.49) | 317 (79.79) | 80 (20.21) | 351 (77.80) | 100 (22.20) |
| Higher | 1 (50.64) | 1 (49.36) | 13 (77.54) | 4 (22.46) | 15 (75.30) | 5 (24.70) |
| **Marital status** | 0.6842 |  | 0.1775 |  | 0.3384 |  |
| Not married | 32 (54.13) | 27 (45.87) | 279 (73.61) | 100 (26.39) | 319 (70.94) | 131 (29.06) |
| Married | 51 (49.94) | 50 (50.07) | 902 (69.09) | 404 (30.91) | 981 (67.93) | 463 (32.07) |
| **Wealth Index of the mother** | 0.9200 |  | 0.0821 |  | 0.1169 |  |
| Poorest | 25 (51.17) | 24 (48.83) | 456 (68.68) | 208 (31.32) | 501 (67.91) | 237 (32.09) |
| Poorer | 27 (56.59) | 21 (43.41) | 407 (75.12) | 135 (24.88) | 440 (73.27) | 160 (26.73) |
| Middle | 21 (50.44) | 20 (49.56) | 224 (64.91) | 121 (35.09) | 256 (63.89) | 145 (36.11) |
| Richer | 7 (44.18) | 8 (55.82) | 66 (65.34) | 35 (34.66) | 73 (62.62) | 43 (37.38) |
| Richest | 3 (41.28) | 5 (58.72) | 27 (86.17) | 4 (13.83) | 32 (77.92) | 9 (22.08) |
| **Mother currently working** | 0.8562 |  | 0.0001* |  | 0.0002* |  |
| No | 41 (50.61) | 40 (49.39) | 678 (75.71) | 218 (24.29) | 739 (73.86) | 261 (26.14) |
| Yes | 41 (52.37) | 38 (47.63) | 503 (63.74) | 286 (36.26) | 562 (62.83) | 333 (37.17) |
| **Frequency of listening to rad** | 0.0098* |  | 0.0307* |  | 0.0057* |  |
| Not at all | 36 (39.56) | 55 (60.44) | 752 (73.80) | 267 (26.20) | 832 (72.95) | 309 (27.05) |
| <once a week | 9 (46.92) | 10 (53.08) | 138 (62.74) | 82 (37.26) | 151 (61.99) | 93 (38.01) |
| ≥ once a week | 13 (73.17) | 5 (26.83) | 143 (65.79) | 74 (34.21) | 151 (63.430) | 87 (36.57) |
| Almost everyday | 20 (60.98) | 14 (39.02) | 148 (64.83) | 81 (35.17) | 166 (61.16) | 106 (38.84) |
| **Frequency of reading NS/MZ** | 0.0581 |  | 0.1561 |  | 0.3137* |  |
| Not at all | 79 (53.57) | 69 (46.43) | 1041 (70.19) | 442 (29.81) | 1153 (68.85) | 522 (31.15) |
| <once a week | 2 (22.60) | 6 (77.40) | 85 (79.36) | 22 (20.64) | 87 (74.51) | 30 (25.49) |
| ≥ once a week | 2 (36.16) | 3 (63.84) | 43 (59.42) | 29 (40.51) | 47 (59.29) | 32 (40.71) |
| Almost everyday |  |  | 12 (54.18) | 10 (45.82) | 13 (56.47) | 10 (43.53) |
| **Frequency of watching television** | 0.4259 |  | 0.8791 |  | 0.9799 |  |
| Not at all | 66 (53.88) | 56 (46.12) | 1006 (69.76) | 436 (30.24) | 1100 (68.57) | 504 (31.43) |
| <once a week | 6 (36.03) | 11 (63.67) | 68 (74.98) | 23 (25.02) | 76 (68.94) | 34 (31.06) |
| ≥ once a week | 1 (18.75) | 4 (81.24) | 55 (70.17) | 24 (29.83) | 57 (67.19) | 28 (32.81) |
| Almost everyday | 10 (60.81) | 6 (39.19) | 52 (70.96) | 21 (29.04) | 67 (70.85) | 28 (29.15) |
| **Sex of child** | 0.1504 |  | 0.0048* |  | 0.0003* |  |
| Male | 45 (47.01) | 51 (52.99) | 599 (66.76) | 298 (33.24) | 658 (64.77) | 358 (35.23) |
| Female | 37 (58.24) | 27 (41.76) | 582 (73.93) | 205 (26.07) | 642 (73.13) | 236 (26.87) |
| **Birth Order** | 0.0004* |  | <0.0001 |  | <0.0001* |  |
| 1-3rd birth order | 37 (63.39) | 21 (36.61) | 682 (86.41) | 107 (13.59) | 741 (85.02) | 130 (14.98) |
| 4-6th birth order | 39 (61.11) | 25 (38.89) | 338 (71.39) | 136 (28.61) | 390 (70.17) | 166 (29.83) |
| 7th + order | 7 (18.85) | 32 (81.15) | 161 (38.18) | 261 (61.82) | 169 (36.27) | 298 (63.73) |
| **Place of Delivery** | 0.3923 |  | 0.015* |  | 0.0187* |  |
| Home | 7 (81.33) | 1 (18.67) | 164 (83.53) | 32 (16.47) | 179 (83.87) | 34 (16.13) |
| Private health facility | 8 (100.00) | 0 (0.00) | 84 (95.45) | 4 (4.55) | 1003 (89.13) | 122 (10.87) |
| Government | 65 (79.26) | 17 (20.74) | 912 (89.91) | 102 (10.09) | 92 (95.83) | 4 (4.17) |
| Unspecified | 4 (100.00) | 0 (0.00) | 21 (82.61) | 4 (17.39) | 27 (81.92) | 36 (18.08) |
| **Birth weight** | 0.0004* |  | <0.0001* |  | <0.0001* |  |
| Less than 2500 | 37 (63.39) | 21 (36.61) | 72 (80.83) | 17 (19.17) | 82 (80.71) | 20 (19.29) |
| 2500 - 3999 | 39 (61.11) | 25 (38.89) | 789 (95.05) | 41 (4.95) | 862 (94.27) | 52 (5.73) |
| 4000+ | 7 (18.85) | 32 (81.15) | 319 (79.00) | 85 (21.00) | 357 (79.02) | 95 (20.98) |
| **ANC visits** | 0.7065 |  | 0.6983 |  | 0.8741 |  |
| <8 ANC visits | 81 (86.97) | 12 (13.02) | 1142 (95.19) | 58 (4.81) | 1257 (94.59) | 72 (5.41) |
| 8+ ANC visits | 1 (100.00) | 0 (0.00) | 17 (92.95) | 1 (7.05) | 19 (93.68) | 1 (6.32) |
| **HIV Results** |  |  |  |  | <0.0001* |  |
| HIV negative | NA | NA | NA | NA | 1180.75 | 503 (29.89) |
| HIV positive | NA | NA | NA | NA | 83 (51.48) | 78 (48.52) |

All tests used the Rao – Scott Chi-square test, ANC: Antenatal Care, NA: Not Applicable

**Table 3:**
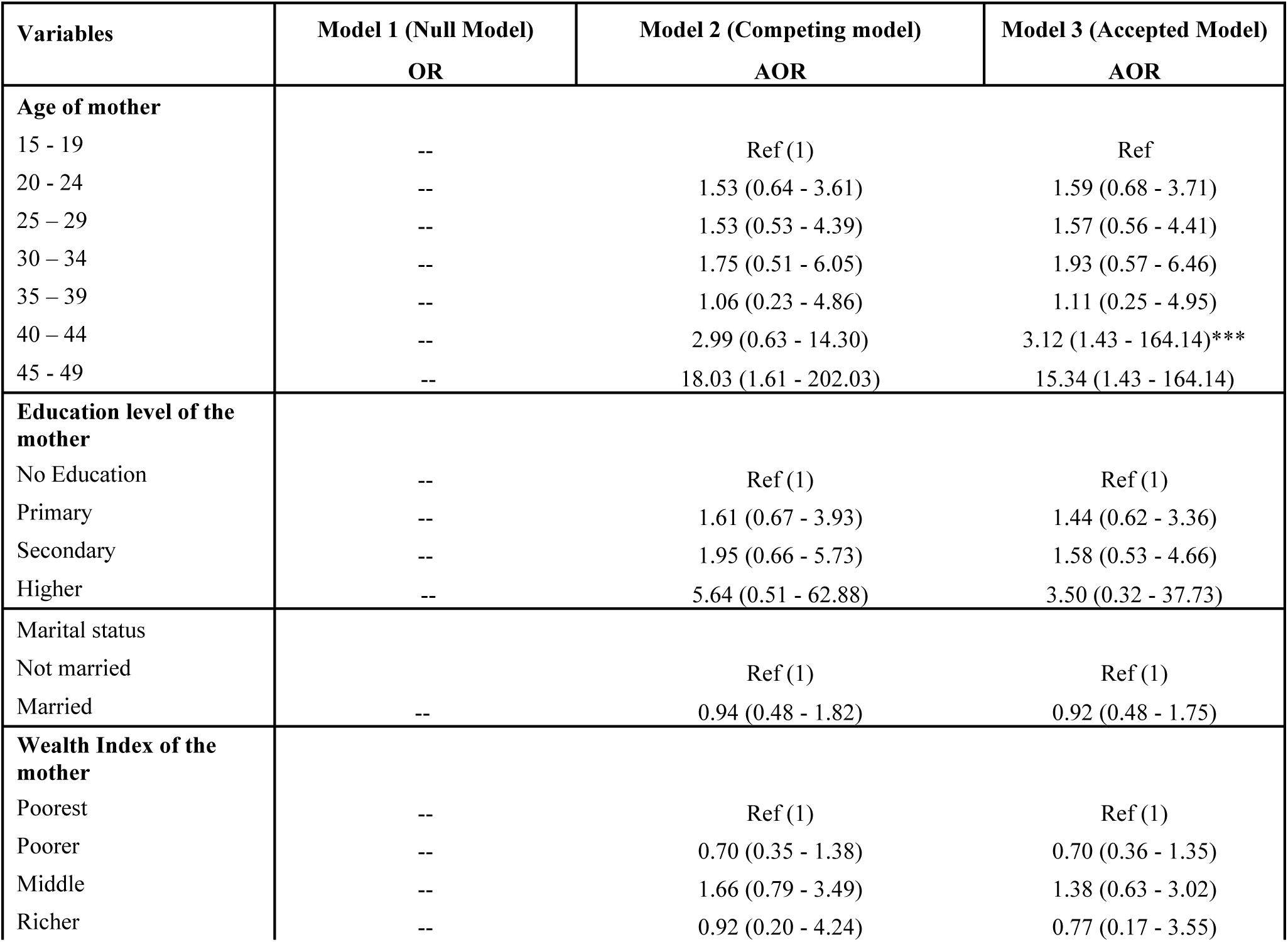
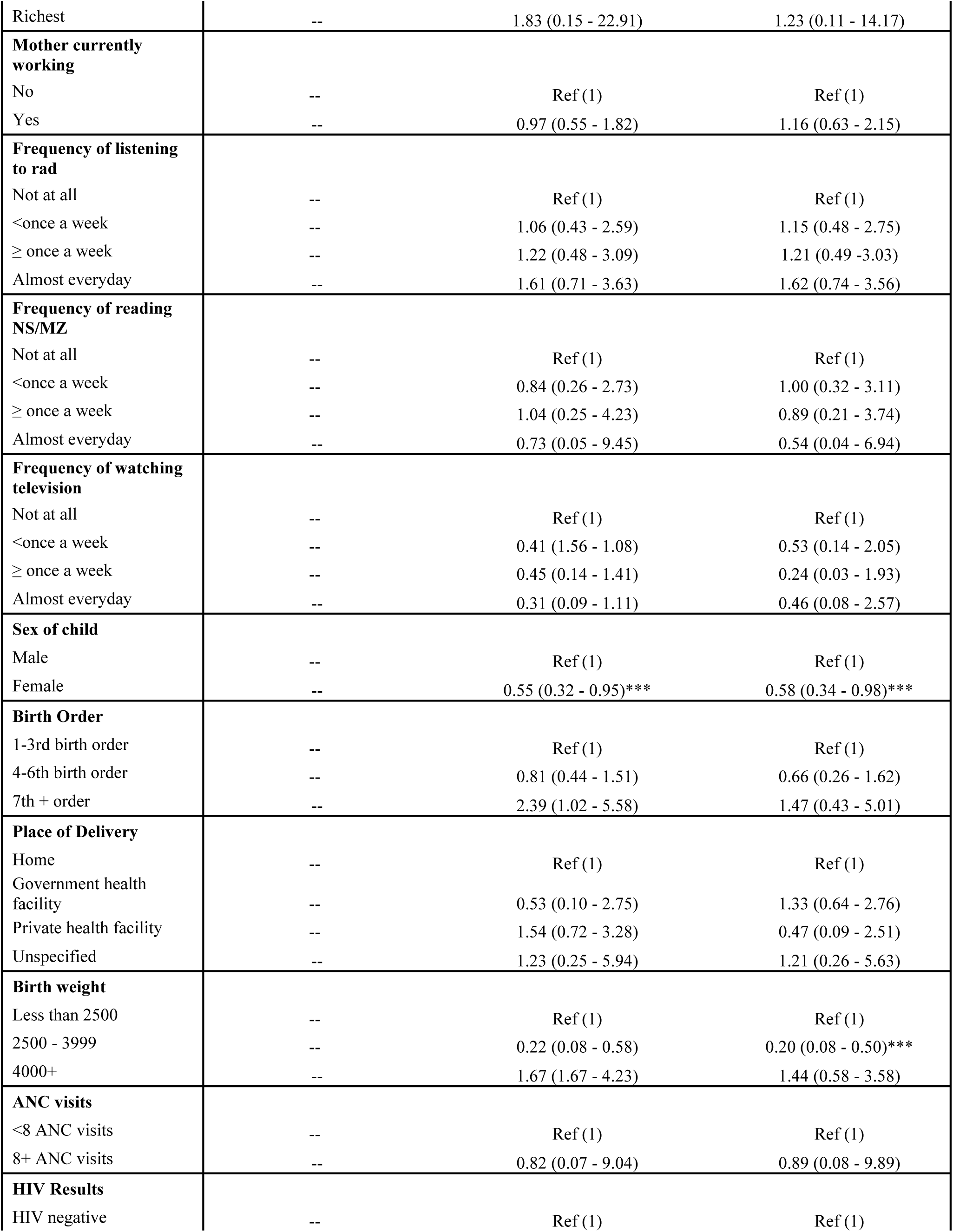
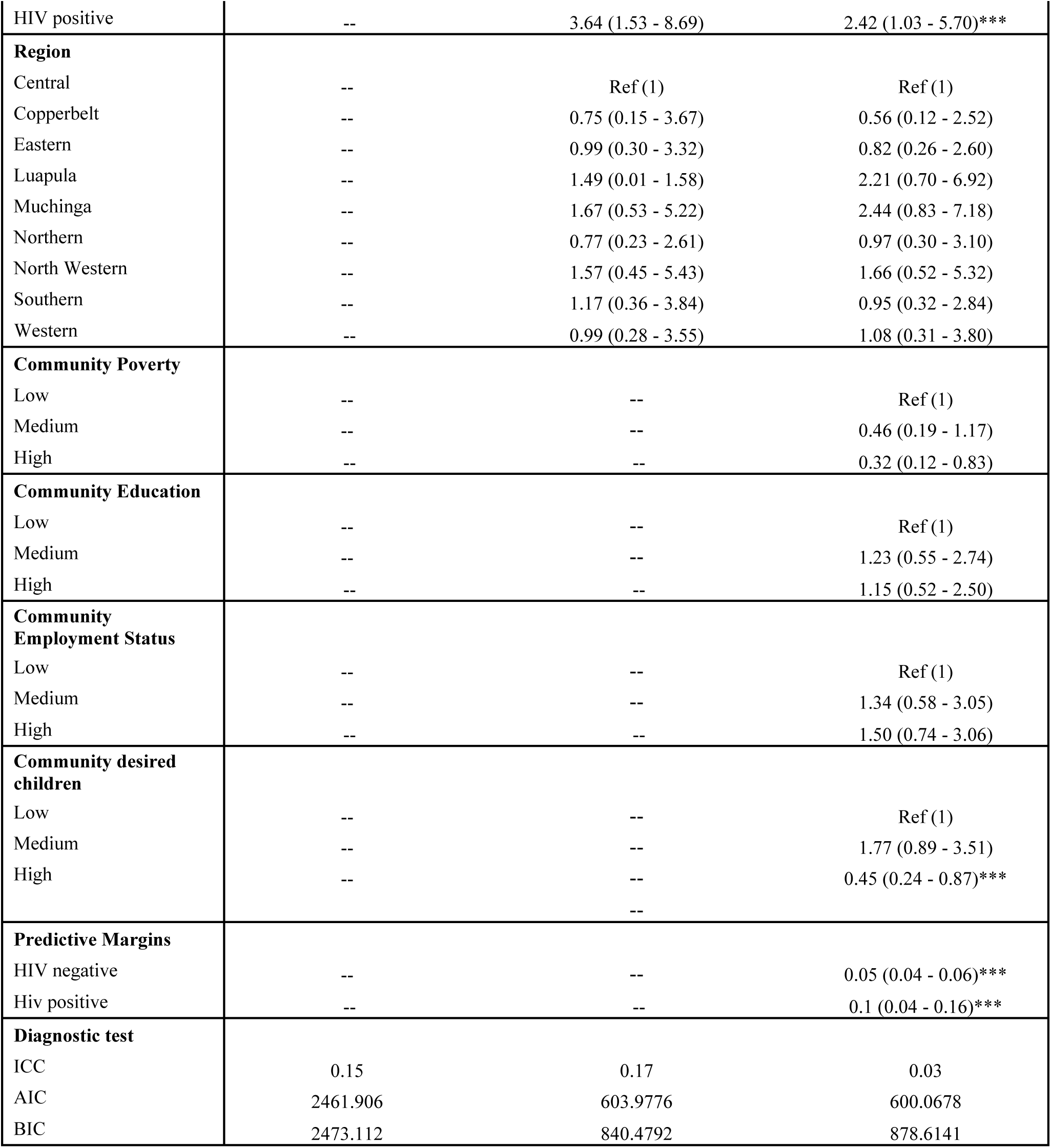
Multilevel mixed effect logistic regression of neonatal mortality of HIV-infected and non-infected mothers in rural Zambia.

Furthermore, mortality rates were significantly elevated among non-HIV-infected mothers lacking formal education (40.78%), compared to 54.56% among HIV-infected mothers. Overall, neonatal mortality stood at 41.13% among mothers (both HIV and non-HIV-infected) with no formal education. Neonatal mortality was significantly higher among HIV-infected mothers who were not working (49.39%), in contrast to non-HIV-infected mothers, where high mortality was observed among those who were employed (36.36%).The neonatal mortality rate was high at 37.17% among neonates whose mothers (both HIV infected and non-infected mothers) were reported to be working. Neonatal mortality was also higher among males in both HIV-infected and non-infected mothers (52.99% and 33.24%, respectively). Moreover, neonatal mortality was consistently higher among children with a birth order of 7 or higher among HIV-infected and non-infected mothers in rural Zambia (81.15% and 61.82%), with overall mortality also reported high among those of the 7th or higher birth order (63.73%).

Additionally, neonatal mortality was reported higher among neonates born with a weight of 4000 grams or higher among HIV-infected and non-infected mothers (81.15% and 21.00%, respectively). Overall, neonatal mortality was higher among mothers infected with HIV (51.48%) compared to those not infected (29.89%).

### Geospatial analysis of neonatal mortality across rural Zambia among HIV-infected mothers

The study examined the geospatial patterns of neonatal mortality among mothers infected with HIV in rural Zambia. It revealed that North-western and Eastern provinces exhibited a high proportion of neonatal mortality, followed by Western and Central provinces. In contrast, Southern and Luapula provinces reported the lowest rates of neonatal mortality among HIV-infected mothers (see Fig 2).

**Figure 2:**
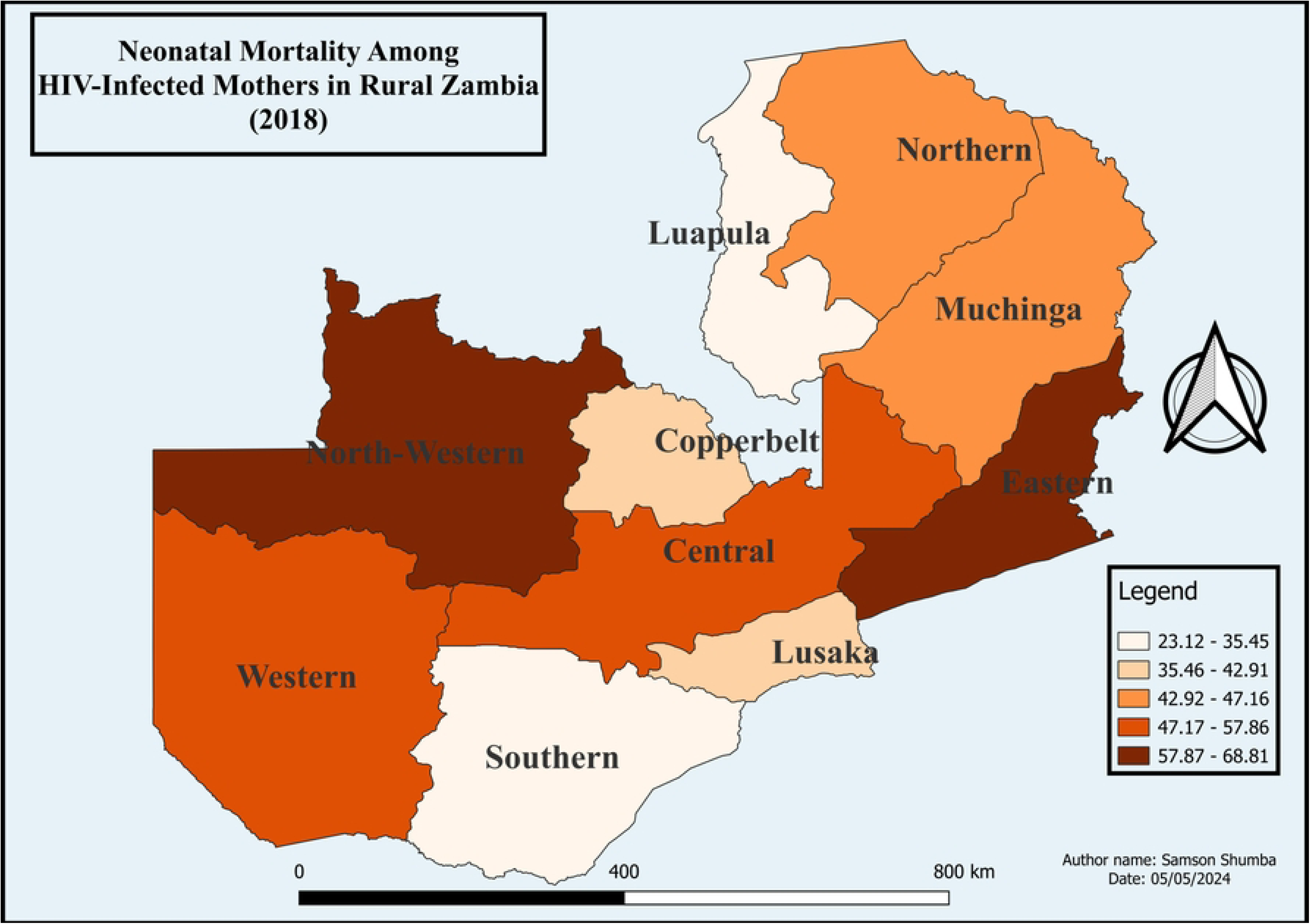
Shows the proportion of neonatal mortality among HIV infected mothers in rural Zambia (2018) The map was generated using QGIS version 3.34.1. (Source: author generated)

**Figure 3:**
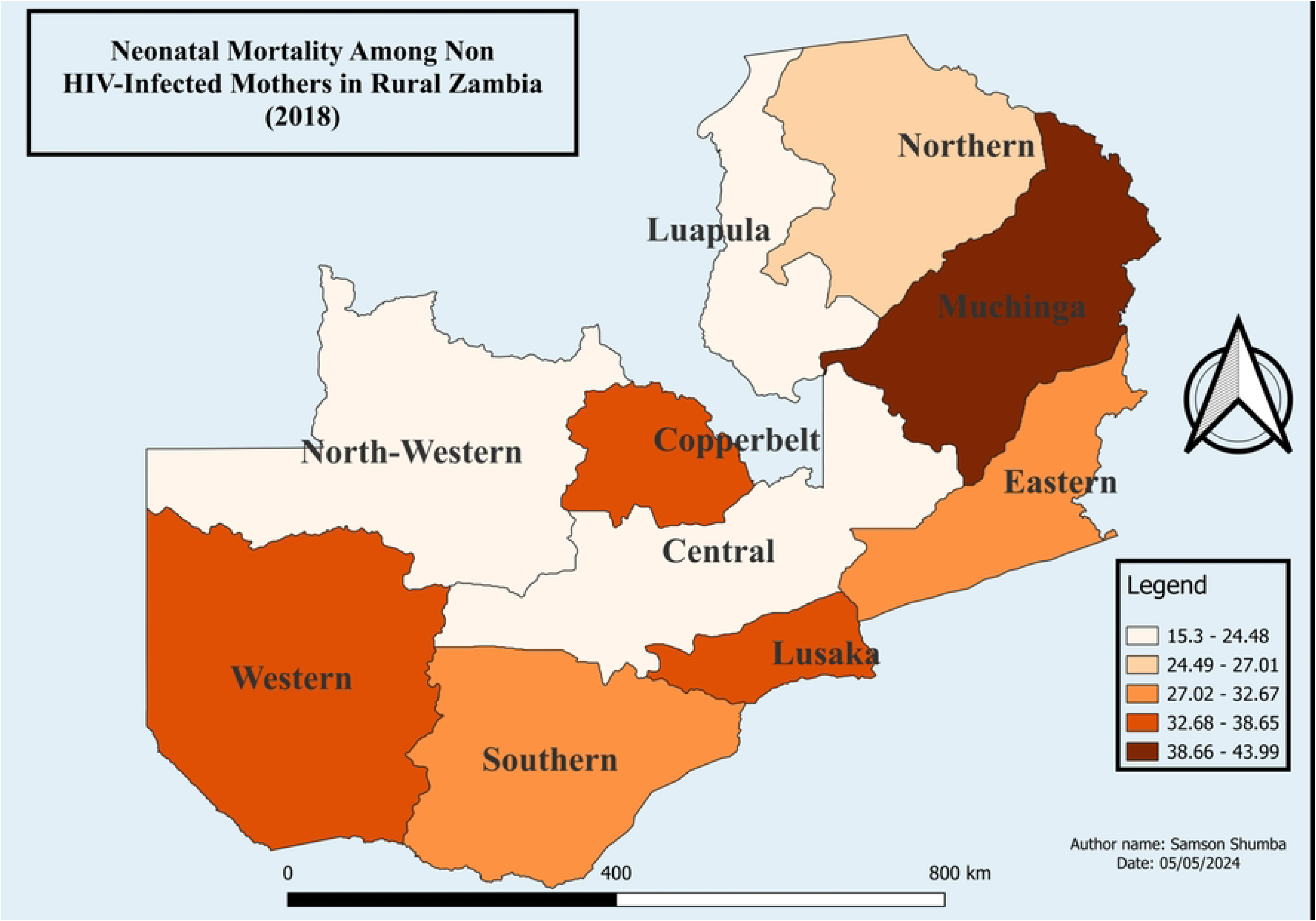
Shows the proportion of neonatal mortality among non HIV-infected mothers in rural Zambia (2018) The map was generated using QGIS version 3.34.1. (Source: author generated)

### Geospatial analysis of neonatal mortality across rural Zambia among non-HIV infected mothers (2018)

The study revealed geospatial patterns of neonatal mortality among non-HIV-infected mothers in rural Zambia. Specifically, Muchinga provinces emerged as a frontrunner in terms of neonatal mortality rates, with Western, Copperbelt and Lusaka provinces following closely behind. Conversely, North-western, Luapula and Central provinces exhibited the lowest rates of neonatal mortality as shown in Fig 4.

**Figure 4:**
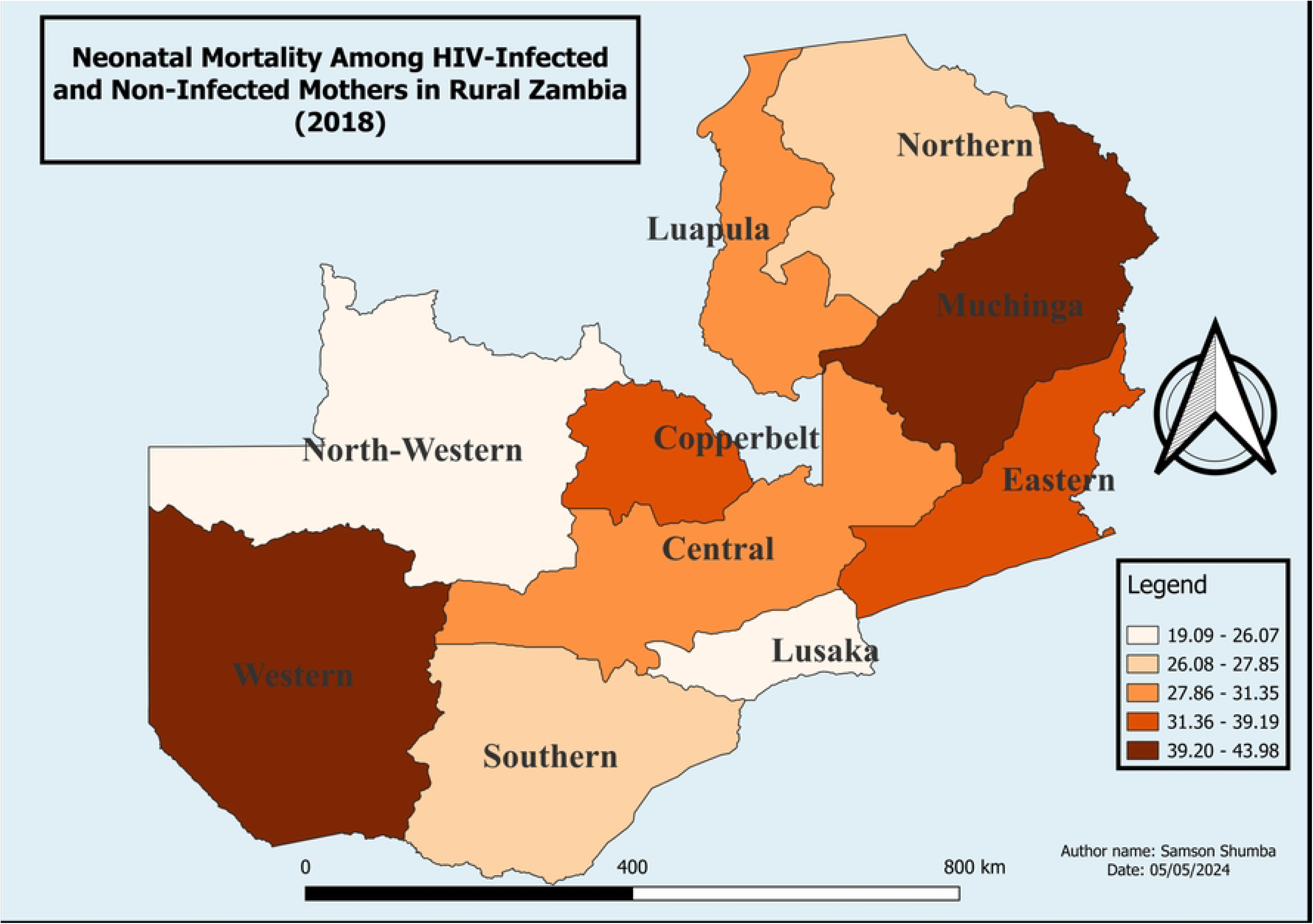
Shows the proportion of neonatal mortality among HIV infected and non-HIV infected mothers in rural Zambia (2018) The map was generated using QGIS version 3.34.1. (Source: author generated)

### Geospatial analysis of neonatal mortality across rural Zambia among HIV infected and non-HIV infected mothers

The study conducted a spatial analysis of neonatal mortality among both HIV-infected and non-infected individuals. The results indicate that neonatal mortality rates were particularly high in Muchinga and Western provinces, with Copperbelt and Eastern provinces following closely behind. Conversely, Lusaka and North-Western provinces reported the lowest proportions of neonatal mortality (see Fig 4).

### Multilevel mixed effect logistic regression of neonatal mortality of HIV-infected and non-infected mothers in rural Zambia

The study employed a multilevel logistic regression model to examine various factors associated with neonatal mortality. Results indicate that mothers aged 40 to 44 years had significantly higher odds of neonatal mortality compared to those aged 15 to 19 years (Adjusted Odds Ratio [AOR]: 3.12, 95% Confidence Interval [CI]: 1.43 – 164.14; p<0.05). Additionally, female neonates exhibited lower odds of mortality compared to males (AOR: 0.58; 95% CI: 0.34 – 0.98; p<0.05). Moreover, neonates with normal birth weight (2500g to 3999g) demonstrated decreased odds of mortality compared to those with low birth weight (less than 2500g) (AOR: 0.2; 95% CI: 0.08 – 0.50; p<0.05). Conversely, neonates born to mothers infected with HIV showed increased odds of mortality compared to those born to HIV-negative mothers (AOR: 2.42, 95% CI: 1.03 – 5.70; p<0.05). Furthermore, the study investigated community-level factors and found that neonates from communities with a higher desired number of children had reduced odds of mortality compared to those from communities with a lower desired number of children holding all factors constant.

The study delved further into examining margin probabilities, revealing that the likelihood of neonatal mortality among HIV-infected mothers in rural Zambia was notably higher compared to non-infected mothers, with probabilities of 0.1 (95% CI, 0.04 – 0.16) and 0.05 (95% CI, 0.04 – 0.06), respectively. The selection of the best-fit model was determined through Akaike Information Criteria (AIC), where Model 3 (Individual plus community level model) was favored over Model 2 (individual level model) and Model 1 (the null model). Additionally, an Intraclass correlation (ICC) of 0.03 was identified, suggesting that merely 3% of the total variance in the outcome variable can be attributed to differences between groups, while the vast majority, 97%, stems from differences within groups. This indicates a low level of clustering or similarity within groups, signifying diverse outcomes within clusters with minimal resemblance among them.

### Predictive margins of neonatal mortality among HIV infected and non-infected mothers in rural Zambia

Additionally, the study presents a graphical depiction of the predictive probabilities of neonatal mortality. The results demonstrate that neonates born to HIV-infected mothers had a significantly higher mortality rate compared to those born to mothers not infected with HIV, as shown in Fig 5.

**Figure 5:**
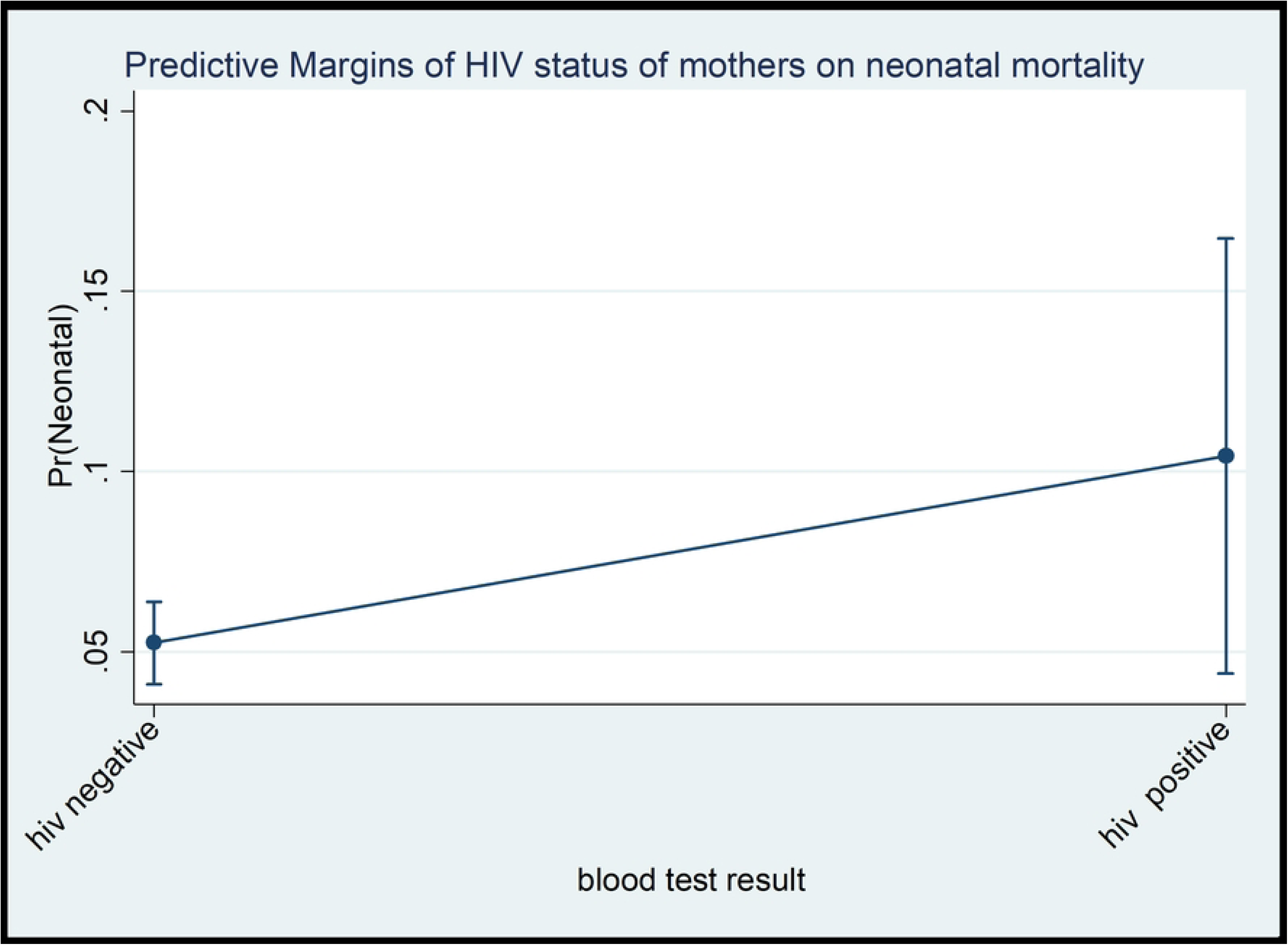
Shows the predictive probabilities of neonatal mortality across rural Zambia among HIV infected and non-HIV infected mothers (2018)

## Discussion

This study investigated the spatial patterns and predictors of neonatal mortality among HIV-infected and non-infected mothers in rural Zambia, utilizing the 2018 Zambia Demographic Health Survey (ZDHS) dataset. Our analysis revealed distinctive patterns in neonatal mortality across various demographic factors shedding light on the intricate interplay of these factors influencing neonatal mortality survival and highlighting regional disparities in neonatal mortality rates in rural Zambia among HIV infected and non-infected mothers. Notably, neonatal mortality exhibited a significant association with an increase in maternal age, reaching its peak in the 45 to 49 age group, accounting for 100% among HIV infected mothers compared to 95.21% in non-HIV-infected mothers. Furthermore, the geospatial findings revealed that the region with the highest rates of neonatal mortality consistently for the three time periods of the demographic health surveys was Western Province; Lusaka, Eastern, and Muchinga fluctuated, and Copperbelt maintained second place throughout the periods.

The study’s findings regarding the association between maternal age and neonatal mortality are consistent with previous research, which has consistently demonstrated a heightened risk of neonatal complications with increasing maternal age [22,23]. This alignment underscores the well-established understanding that rates of adverse pregnancy outcomes, including neonatal mortality, tend to rise significantly as maternal age advances [24]. These consistent findings emphasize the crucial importance of educating women about the potential risks associated with delayed motherhood [25]. Conversely, a study in Brazil focusing solely on the impact of maternal age on neonatal death found elevated mortality rates among adolescent mothers compared to their older counterparts [26]. However, our study, employing multilevel logistic regression, revealed that women aged 45 to 49, regardless of HIV infection status, exhibited increased odds of neonatal mortality compared to mothers aged 15 to 19 years. This finding contrasts with research conducted in Bangladesh, where women aged 19 years and above showed higher odds of neonatal mortality compared to those younger than 19 [27–29].

Furthermore, our investigation revealed that female neonates exhibited a lower likelihood of neonatal mortality, indicating that male neonates were more vulnerable to mortality within the first month of life compared to their female counterparts. These findings align with prior research indicating a higher mortality rate among boys compared to girls [30,31]. Additionally, our results indicated that neonates with normal birth weight had a decreased likelihood of mortality compared to those with low birth weight, consistent with studies conducted in Kenya [32] and Nepal [33].

Furthermore, our study emphasized maternal HIV status on neonatal mortality in rural Zambia. The analysis of predictive margins revealed that neonates born to HIV-infected mothers faced an increased probability of mortality compared to those born to non-infected mothers. Multiple studies have highlighted alarmingly high mortality rates among HIV-exposed children [34,35]. Studies have shown that HIV exposure is associated with low birth weight, prematurity, and a higher proportion of infants being small for gestational age [36–38]. Moreover, chronic immune activation resulting from maternal HIV infection impacts infant immunology levels, and HIV-exposed but uninfected children often possess lower levels of maternally transferred antibodies at birth [35].

Our study investigated geospatial patterns of neonatal mortality among HIV-infected and non-infected mothers of rural Zambia across 10 provinces of Zambia. Nationally, neonatal mortality levels exhibited spatial variation, with hotspots identified in North-western and Eastern provinces among HIV-infected mothers, and Muchinga province among non-HIV-infected mothers. This discrepancy in neonatal mortality rates across provinces may stem from multiple factors, including disparities in access to healthcare services, variations in socio-economic conditions, differences in maternal education levels, prevalence of diseases, and cultural practices related to childbirth and infant care. The data emphasizes the dynamic nature of neonatal mortality proportions across Zambia’s provinces during the specified period. Understanding the underlying factors contributing to these regional disparities is paramount for formulating targeted and effective interventions aimed at reducing neonatal mortality rates and improving maternal and child health outcomes nationwide.

The Sustainable Development Goals (SDGs) have set a global target to reduce neonatal mortality rates to no more than 12 infant deaths per 1,000 live births by 2030 [39]. However, achieving this target faces significant challenges, particularly in regions such as sub-Saharan Africa, where numerous factors contribute to elevated neonatal mortality levels. According to UNICEF, a lot of countries are at risk of missing the SDG target on neonatal mortality than on under-five mortality [40]. Moreover, our study findings emphasizes the multifaceted nature of factors influencing neonatal mortality, including maternal age, neonate sex, birthweight, maternal HIV status, and community desired number of children. This intricate web of variables highlights the need for a shift in focus within current mother and child health programs to incorporate activities that empower women to make informed decisions regarding the timing of childbirth, monitor their HIV status, consider the birthweight and sex of their child, among other crucial factors. Moreover, a systematic review study and meta-analysis conducted in sub-Saharan Africa revealed that rural residence was a significant contributing factor to high neonatal mortality rates [41]. Our study’s novel findings offer valuable insights previously unexplored, illuminating the intricate landscape of neonatal mortality in rural Zambia. Recognizing the distinct patterns of neonatal mortality between HIV-infected and non-infected mothers is crucial for tailored interventions and enhancing maternal and child health outcomes within these communities.

### Study strength and study Limitations

This study benefits from the utilization of national data, offering a representative sample of neonates, thus enabling generalizability to this specific demographic. However, certain limitations must be acknowledged. The study relies on the 2018 Zambia Demographic and Health Survey (ZDHS) dataset, employing a cross-sectional design, which indicates correlation rather than causation between the outcome and individual or contextual factors. Additionally, the utilization of contextual and community-level factors from the ZDHS may not fully capture the community experience. Moreover, potential recall bias exists, with mothers possibly misreporting crucial information like their child’s age at death, birth order, and birth weight. To address this, rigorous data cleaning procedures were implemented to identify and rectify inconsistencies or outliers.

### Policy recommendation

Zambia has demonstrated a steadfast commitment to improving neonatal survival through targeted interventions from 2000 to 2017, focusing on bolstering healthcare infrastructure, community outreach programs, and healthcare worker capacity building. Fluctuations in neonatal mortality across provinces are likely influenced by the implementation of key programs like maternal and perinatal death reviews, essential newborn care, and Kangaroo mother care. To address rural Zambia’s neonatal mortality, a comprehensive approach is recommended. This includes strengthening healthcare infrastructure, enhancing community outreach, providing ongoing training for healthcare workers, improving maternal education, and tailoring interventions to address regional disparities based on factors like access to healthcare and socio-economic conditions. These efforts signify Zambia’s dedication to improving maternal and child health outcomes and enhancing neonatal survival rates.

## Conclusion

In conclusion, the findings in this study provide valuable insights into the spatial patterns and determinants of neonatal mortality among HIV-infected and non-infected mothers in rural Zambia. The findings reveal distinct patterns of neonatal mortality across various demographic factors, emphasizing the complex interplay of these factors and highlighting regional disparities in neonatal mortality rates. Notably, maternal age, sex of the neonate, birthweight, HIV status of the mother, and community’s desired number of children emerged as significant predictors of neonatal mortality. Geospatial analysis identified hotspots of neonatal mortality in certain provinces, underscoring the need for targeted interventions tailored to regional contexts. Furthermore, the study emphasizes the need for strengthening healthcare infrastructure, promoting community outreach programs, enhancing healthcare worker training, focusing on maternal education, and addressing regional disparities. These recommendations aim to improve maternal and child health outcomes and contribute to the achievement of Sustainable Development Goals (SDGs) targets.

## Data Availability

Authorization to utilize the data was secured from ICF Macro, accessible at https://dhsprogram.com/data, under the dataset titled ZMKR71FL.DTA and ZMAR71FL.DTA

## Acknowledgement

Many thanks to the Zambia Statistical Agency (ZSA) and the DHS program for granting permission to utilize the 2018 ZDHS.

## Dedication

This paper is dedicated, with love and reverence, to the cherished memory of Wise Mwila Katumbo.

